# Seizure Forecasting Using a Novel Sub-Scalp Ultra-Long Term EEG Monitoring System

**DOI:** 10.1101/2021.05.09.21256558

**Authors:** RE Stirling, PJ Karoly, MI Maturana, ES Nurse, K McCutcheon, DB Grayden, SG Ringo, J Heasman, TL Cameron, RJ Hoare, A Lai, W D’Souza, U Seneviratne, L Seiderer, KJ McLean, KJ Bulluss, M Murphy, BH Brinkmann, MP Richardson, DR Freestone, MJ Cook

## Abstract

Accurate identification of seizure activity, both clinical and subclinical, has important implications in the management of epilepsy. Accurate recognition of seizure activity is essential for diagnostic, management and forecasting purposes, but patient-reported seizures have been shown to be unreliable. Earlier work has revealed accurate capture of electrographic seizures and forecasting is possible with an implantable intracranial device, but less invasive electroencephalography (EEG) recording systems would be optimal. Here, we present preliminary results of seizure detection and forecasting with a minimally invasive sub-scalp device that continuously records EEG.

Five participants with refractory epilepsy who experience at least two clinically identifiable seizures monthly have been implanted with sub-scalp devices (Minder™), providing two channels of data from both hemispheres of the brain. Data is continuously captured via a behind-the-ear system, which also powers the device, and transferred wirelessly to a mobile phone, from where it is accessible remotely via cloud storage. EEG recordings from the sub-scalp device were compared to data recorded from a conventional system during a 1-week ambulatory video-EEG monitoring session. Suspect epileptiform activity (EA) was detected using machine learning algorithms and reviewed by trained neurophysiologists. Seizure forecasting was demonstrated retrospectively by utilising cycles in EA and previous seizure times.

The procedures and devices were well tolerated, and no significant complications have been reported. Seizures were accurately identified on the sub-scalp system, as confirmed by periods of concurrent conventional scalp EEG recordings. The data acquired also allowed seizure forecasting to be successfully undertaken. The area under the receiver operating characteristic curve (AUC score) achieved (0.88) is comparable to the best score in recent, state-of-the-art forecasting work using intracranial EEG.

## Introduction

For people with epilepsy, an estimation of total seizure burden is fundamental to clinical management as well as for the evaluation of new therapies, such as drugs or devices. For over a century, clinicians have relied on their patients’ reports of their seizure frequency, “that it may be taken as an index of the severity of the epileptic condition”(1). Although the rate of clinical seizures influences an individual’s perception of disease severity, the physiological basis for this remains ambiguous (2,3). Indeed, the number of clinical seizures is not representative of (nor closely correlated with) the total seizure burden (4). Rates of subclinical epileptiform activity seen on electroencephalography (EEG) are typically orders of magnitude higher than clinical seizures. These subclinical events may impact cognition (5,6) and quality of life, and are important in epilepsy diagnosis and treatment, particularly for syndromes that are characterised by stereotypical discharges. Epileptiform activity (EA) is also relevant for surgical planning (7) and forecasting seizure likelihood (8). Hence, capturing both clinical and subclinical EA is important for the clinical management of epilepsy.

The easiest and most common method of capturing clinical EA is through patient self-reporting. Unfortunately, the accuracy of self-reported EA is unreliable (9,10). In addition to unawareness of subclinical events, patients are often unaware or amnestic for their clinical seizures and may also report other non-epileptic symptoms as seizures. As there have been no real alternatives, seizure diaries (both paper and electronic) are used almost exclusively to manage patients, and regulatory authorities assess new treatments primarily on evidence from diaries (11). It is possible that the unreliability of self-reporting has impeded progress in the development of anti-seizure medications (12). In addition to inaccurate records of seizure frequency, people with epilepsy and caregivers typically cannot provide an objective assessment of the time of seizure onset, seizure duration or seizure type (11). This detailed information about seizures is important for patient management, particularly with regard to medication titration and safety. For this reason, capturing EEG correlates of seizures remains the reference standard in clinical epilepsy management.

Short-term (up to 10 days) inpatient video-EEG assessment can be used to assess treatment efficacy, for surgical planning, and has been proposed as an objective metric for randomized controlled trials. However, short-term monitoring has major limitations. The spatiotemporal organization of subclinical EA, including epileptiform spikes and high frequency oscillations (HFOs), changes over long time scales (months to years), so short-term capture of subclinical EA is unreliable (13,14). In addition, seizure rates show high natural variability and require long-term recording to identify clinically relevant improvements (15,16). Short-term monitoring is particularly inadequate for people with lower seizure frequencies and cannot detect multiday cycles of EA that occur in most individuals (17,18).

Ultra-long term monitoring is required for better diagnosis, management, and treatment of epilepsy, including seizure forecasting. Currently, scalp EEG is not suitable for ultra-long term monitoring due to limited data quality and the need for external electrode maintenance (19). Invasive intracranial systems, such as the RNS System (NeuroPace) and the Percept PC (Medtronic), are available but are built for neurostimulation, do not store sufficient data and are too invasive for diagnostic applications (19). Alternatively, sub-scalp EEG systems are minimally-invasive tools that may address the need for objective ultra-long term EEG recordings (19,20), allowing for personalised and accurate epilepsy management.

Our earlier work with an implantable intracranial device (4) demonstrated that continuous EEG permitted characterisation of EEG features (21,22), epileptic activity (18,23) and sleep (24,25), and enabled successful seizure forecasting (26–28). As similar data could be acquired from a less invasive (sub-scalp) EEG recording system, we have developed a minimally invasive device that is inserted into a sub-scalp location to continuously record EEG. This work reports on the feasibility of the system to detect interictal EA and seizures in five subjects. We also demonstrate in one of the participants that seizure forecasting is possible with the sub-scalp device. This case study demonstrates how cycles can be derived from events in the EEG and in turn, these cycles can be used to forecast epileptic seizures. The forecasting method has been built on previous work in seizure cycles (20,29–31) and interictal EA cycles (17,18,32).

## Materials and Methods

### Patient selection and criteria

Data used in this work were acquired during a registered trial (ACTRN 12619001587190). Subjects participating in the Minder™ sub-scalp system (Table 1) trial were 18-75 years of age, had an established clinical diagnosis of epilepsy (33) with a minimum of two clinically identifiable epileptic seizure events per month, and otherwise were medically and neurologically stable as defined by their clinician. All participants had EEG profiles that were consistent with epilepsy diagnosis and had prior neuroimaging. Subjects were excluded if they had a neurostimulation implant device for epilepsy or another condition or had any other condition that may impact the study outcome or safety of the device.

**Table 1.** Participant demographics.

| Participant | Gender | Age (years) | Epilepsy type | Epilepsy aetiology | Antiepileptic medications |
| --- | --- | --- | --- | --- | --- |
| 1 | F | 50-54 | Multifocal | Periventricular nodular heterotopia | vimpat, epilim, lyrica, briviact |
| 2 | M | 60-64 | Focal | Cortical dysplasia | tegretol, vimpat |
| 4 | F | 50-54 | Focal | Hypothalamic hamartoma | tegretol, briviact, phenobarb, rivotril |
| 5 | F | 45-49 | Focal | Non-lesional temporal lobe epilepsy | lamotrigine |
| 6 | M | 45-49 | Generalized | Genetic generalized epilepsy | epilim, keppra, lamictal, zonegran |

All participants wore the sub-scalp system for at least 8 months during both wake and sleep. Subjects were also expected to maintain a seizure diary, if necessary, with the assistance of a caregiver, and attend regular study appointments. All participants gave written, informed consent and the study protocol was approved by St Vincent’s Hospital Melbourne Human Research Ethics Committee (HREC 063/15).

### Implantable System

The Minder™ sub-scalp system (Epi-Minder Pty Ltd) is an investigational device comprising an implanted device, which communicates with an external wearable unit, a mobile phone, and a secure cloud (Figure 1). The implanted device is positioned under the scalp, with a small burr hole to recess the telemetry device and includes an electrode array that are passed superiorly with two contacts located over each parietal bone. The electrodes record differential EEG signals across two contacts at 250 Hz, which are captured by the telemetry unit. The telemetry unit communicates with an external behind-the-ear (BTE) processor via an inductive radio frequency (RF) link, which allows data and power transfer between the external wearable device and the implant. The BTE processor communicates with a mobile phone via Bluetooth. The mobile phone application (Minder app) facilitates the transfer of EEG data from the implant to the phone, and ultimately to a secure cloud for processing. The Minder app also captures audio and accelerometery data from the phone and stores it together with the EEG data in the Seer Cloud (Seer Medical Pty Ltd). Data captured by the implanted device is reviewed and curated on the Seer Cloud platform. Curated events are used for training a machine learning algorithm that detects epileptiform events and whose output is used for seizure forecasting. In future, seizure forecasting will be delivered to patients through the Seer App.

**Figure 1:**
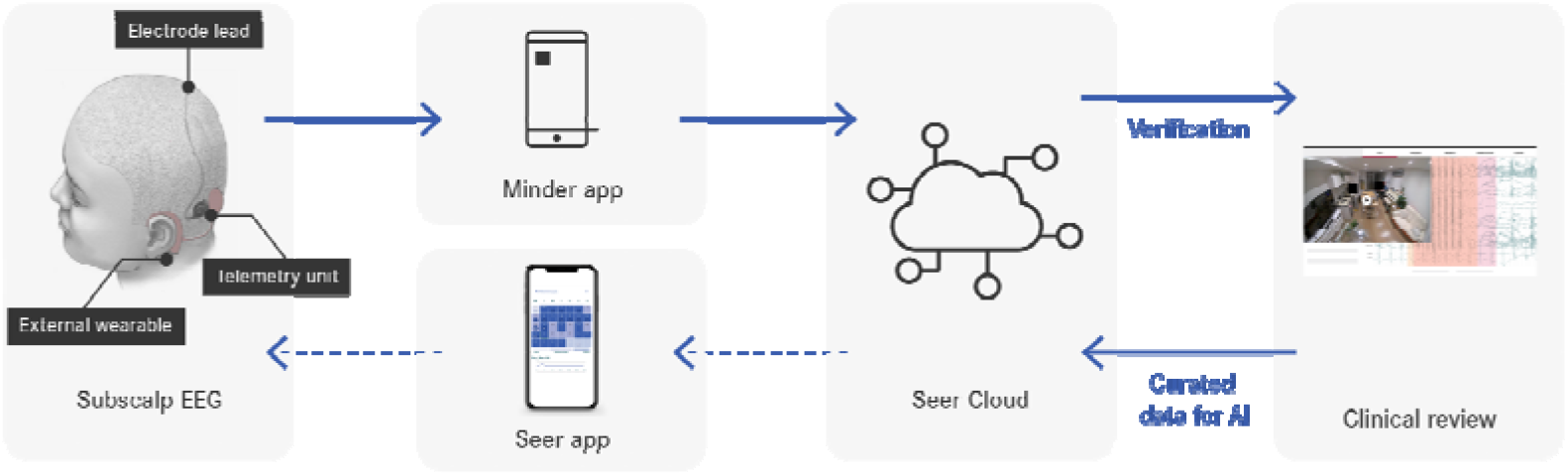
Data flow throughout the system. Dotted arrows indicate system components under development.

### Surgical procedure and follow up assessments

The device is implanted in a specific position for the implanted receiver placed in the mastoid bone and electrode array passed subcutaneously and over the pericranium posterior to the vertex and over the parietal regions, under general anaesthetic. The location of the sub-scalp electrode was chosen to optimise EA detection rates and minimise artifact from nearby muscles. The surgical procedure for implantation of the Minder implant housing and coil was modelled closely on that used for commercial cochlear implants. Regular check-ups (every 2-6 weeks) were conducted in person and participants communicated with study doctors and coordinators between in-person visits. In addition, 7-day scalp EEG was performed at weeks 4 and 24 after implantation. The scalp EEG recordings consisted of a standard 10-20 electrode placement with an additional four scalp electrodes placed as close as practically feasible to the underlying implanted sub-scalp electrodes. The purpose of the scalp EEG assessment was to compare the sub-scalp EEG signal to the scalp EEG, particularly during seizures and interictal discharges, and as well activities including sleep, and potential sources of artefact. Subjects were asked to keep their seizure diaries during monitoring so that the three modalities of seizure detection (sub-scalp implant, scalp EEG and seizure diary) could be compared.

### Epileptiform activity detection and annotation

EA detection was aided by a machine learning algorithm designed to detect relevant events in the EEG (34,35). The algorithm was designed to label suspect EA with high sensitivity to ensure that all events were detected and could be reviewed. EEG recordings and computer generated annotations were accessed from the cloud through an online portal and reviewed by trained neurophysiologists who marked interictal events and seizures. Interictal events consisted of typical epileptiform EEG activity such as spike discharges. EEG seizures consisted of EEG activity substantially larger than background and lasting a minimum of 10 seconds. Seizure morphologies were first confirmed in each participant by observing correlated seizures in the scalp EEG.

### Seizure forecasting case study

This retrospective case study was designed using training and testing datasets. We utilized cycles in both EA and seizures to forecast seizure likelihood per hour. The forecaster attempted to predict confirmed seizure events.

#### Data pre-processing and feature extraction

Two features were incorporated into the forecaster: significant cycles based on rates of machine-detected EA and significant cycles based on seizures. To compute cycles in EA, we used a similar approach to a previously published method for extracting rhythms of EA (17). Briefly, a Morlet wavelet transform was computed on the z-standardised hourly event rates to produce a global wavelet spectrum of power for each scale (cycle period). The cycle periods considered were every 1.2 hours between 2.4 and 31.2 hours, every 2.4 hours between 33.6 and 48 hours, every 4.8 hours between 52.8 and 4 days and every 12 hours between 5 days and up to a maximum period of a quarter of the recording duration. At least four cycle periods had to be present to confirm a cycle. Peaks in the wavelet spectrum were found by comparing neighbouring values. Peaks above the global significance (99% confidence) level were determined to be significant EA cycle periods using a time-averaged significance test (36).

Once significant cycle periods were computed, event rates were filtered at each significant cycle period using a zero-order Butterworth bandpass filter. The bandpass filter used cut-off frequencies at ±33% of the cycle frequency (consistent with (17)). These cut-off frequencies were chosen to account for phase shifts in the cycle over the recording time. To account for bandpass overlap in significant cycle frequencies, we introduced a sparsity criterion whereby only the strongest peak (greatest power in the wavelet spectrum) within any cycle’s bandpass filter pass band was considered. The instantaneous phase of the cycle at each timepoint was then estimated using a Hilbert transform. Filtered cycles in EA rates were used as features for the forecaster if seizures were significantly phase-locked to the cycle (p<0.05, according to the omnibus/Hodges-Ajne test for circular uniformity (37)).

Cycles in seizure times were detected using a similar approach to our previous work (29,38). We assessed the phase locking of seizure times to a range of possible cycles using both the Omnibus test (p < 0.05) and the synchronisation index (SI ≥ 0.4) value to quantify phase locking. The SI value - a measure of the magnitude of the resultant vector - ranges from 0 to 1, where 0 represents a perfectly uniform circular distribution and 1 represents perfect alignment with respect to an underlying cycle (30). To account for multiple cycle periods meeting the criteria within close proximity, we used only the strongest cycle period (based on the highest SI value) within ±33% of any other cycle period.

All features were transformed from cyclical to linear features by normalizing the signals from 0 to 2π and computing the sine and cosine of the normalized signal.

#### Forecasting algorithm

To forecast the likelihood of a seizure on an hourly basis, we used an ensemble machine learning algorithm that combined a random forest (RF) regressor and a logistic regression (LR) classifier. The output of the model was the final likelihood of a seizure (risk value), which was represented as a continuous value between 0 for no seizure and 1 for a guaranteed seizure within the next hour.

The RF regressor with the bootstrap aggregating technique was trained on all features. In the model, the number of decision trees was 80 and the minimum number of samples required to be at a leaf node was 15. From observation, these model parameters achieved the highest accuracy on the training dataset. Since seizures typically account for less than 1% of daily life, the dataset is usually imbalanced, with non-seizure hours occurring far more frequently than seizure hours. RF models typically performed better on balanced datasets, so oversampling of seizure hours was undertaken before training the RF model. The output of the RF model was used as an input to the LR classifier. The LR classifier was trained on all features, including the output of the RF model. For simplicity, the default logistic regression model was used from Python’s *sklearn* library. The output of the LR model was the final likelihood of a seizure (risk value) within the next hour.

Using the likelihood values, the forecaster classified hours as either low, medium, or high risk. The medium and high risk cut-off thresholds were computed by optimising:

(C1) time spent in low risk > time spent in medium risk > time spent in high risk;

C2) seizures in high risk > seizures in medium risk > seizures in low risk;

If C1 or C2 could not be satisfied, the optimisation algorithm maximised the product of the time in low risk and the number of seizures in high risk (C3 and C4):

(C3) maximise the time spent in a low risk state;

(C4) maximise the number of seizures occurring in the high risk state.

Note that the likelihood is distinct from a traditional probability value where all outcomes sum to 1. This distinction is caused by oversampling the seizure class in the RF model, which generates synthetic seizure-hours such that the number of seizure hours is equal to the number of non-seizure hours. The result is that the likelihood values are higher than the true probability values.

#### Training and testing datasets

After pre-processing and feature extraction, the dataset was split into training and testing datasets. Initial algorithm training occurred using seizures captured over the first 14 days (15 seizures) but using cycles derived from the entire dataset. After the initial training, re-training occurred after each new seizure was observed. Re-training occurred on all past data, which recomputed the algorithm coefficients and risk thresholds for future predictions.

All analyses were executed in Python (version 3.7.9) using pandas (v1.2.0), numpy (v1.19.2), matplotlib (v3.3.2), datetime (v3.7.9), scipy (v1.5.2), pycwt (v0.3.0), sklearn (0.23.1), imblearn (v0.6.2) and pycircstat (v0.0.2) libraries.

## Results

The surgical procedure and devices were well tolerated. No significant complications have been reported in the five participants. The 7-day EEG recordings revealed interictal events in all participants and seizures in four of the five participants (Table 2). Overnight use of the system was well tolerated and the BTE processor was worn either on the ear or attached to the clothing during sleep. Seizures were identified on the sub-scalp system, as confirmed by periods of concurrent conventional scalp EEG recordings (Figure 2A). A correlation between the sub-scalp and scalp EEG recordings during seizures revealed significant correspondence between the sub-scalp and scalp EEG.

**Table 2:** Clinically relevant EEG events during the two 7-day EEG sessions.

| Participant | Interictal events | Seizures | Patient reported seizures (confirmed events) |
| --- | --- | --- | --- |
| 1 | S1: 1476<br>S2: 5981 | S1: 27<br>S2: 17 | S1: 7 (6)<br>S2: 1 (1) |
| 2* | S1: 245 | S1: 3 | S1: 6 (0) |
| 4 | S1: 179<br>S2: 52 | S1: 3<br>S2: 0 | S1: 4 (3)<br>S2: 0 |
| 5 | S1: 519<br>S2: 110 | S1: 0<br>S2: 0 | S1: 3 (0)<br>S2: 0 |
| 6 | S1: 5783<br>S2: 2084 | S1: 5<br>S2: 0 | S1: 1 (0)<br>S2: 0 |

Confirmed events are seizures confirmed through clinical review
S1 - 7-day monitoring at 4 weeks post-implant
S2 - 7-day monitoring at 24 weeks post-implant
\*: note participant 2 did not have monitoring at 24 weeks

**Figure 2:**
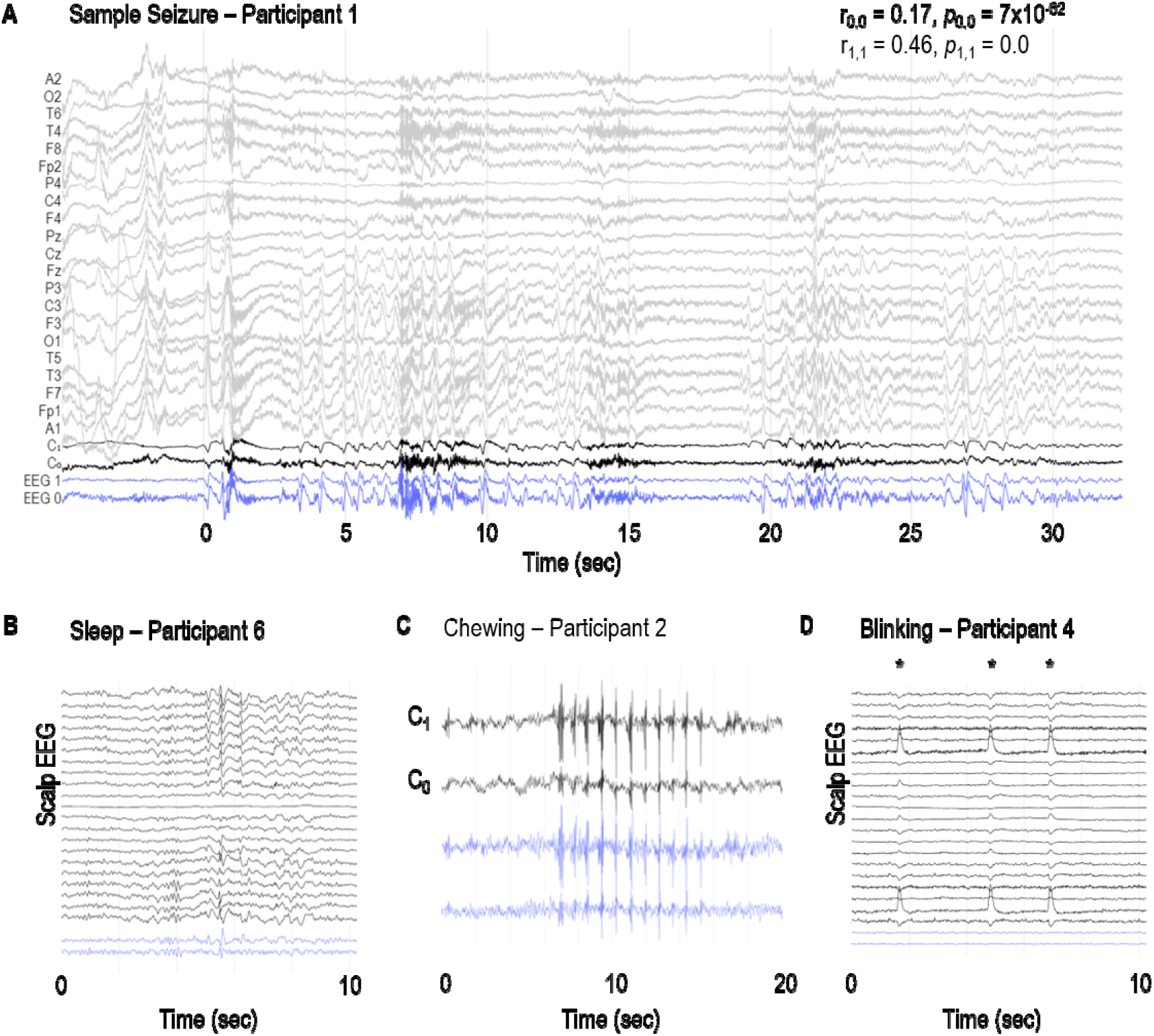
Sample EEG recordings. **(A)** Sample seizure in participant 1 and examples of **(B)** vertex waves and spindles during sleep, **(C)** chewing and **(D)** blinking artefacts (*) in participants 6, 2 and 4, respectively. C_0_ and C_1_ channels represent the bipolar recordings from the additional scalp electrodes placed over the sub-scalp electrodes. Blue traces (EEG 0 and EEG 1) represent the sub-scalp recordings. r_0,0_ and r_1,1_ represent the Pearson correlation coefficient between C_0_ and EEG 0, and C_1_ and EEG 1, respectively. p_0,0_ and p_1,1_ represent the respective p values.

Many other neurological events and artefacts were also present in the sub-scalp recordings. In all participants, clear sleep-related transients were visible in the sub-scalp recordings (Figure 2B). Head scratching and muscular artefacts, such as chewing or jaw clenching artefacts, typically appeared very large across the sub-scalp recordings (Figure 2C), while other artefacts, such as blinking, were largely invisible (Figure 2D).

### Seizure forecasting case study

We conducted a proof-of-principle analysis of seizure forecasting for Participant 1. This participant had a total of 134 seizures over a 6 month period. Figure 3A shows the hourly rate of detected events from a machine learning algorithm over the 6 month period. The machine learning algorithm detects events most similar to interictal spikes. Shaded blue regions represent the two 7-day EEG sessions recorded at weeks 4 and 24. The purple region represents an extended period where data was not collected.

**Figure 3:**
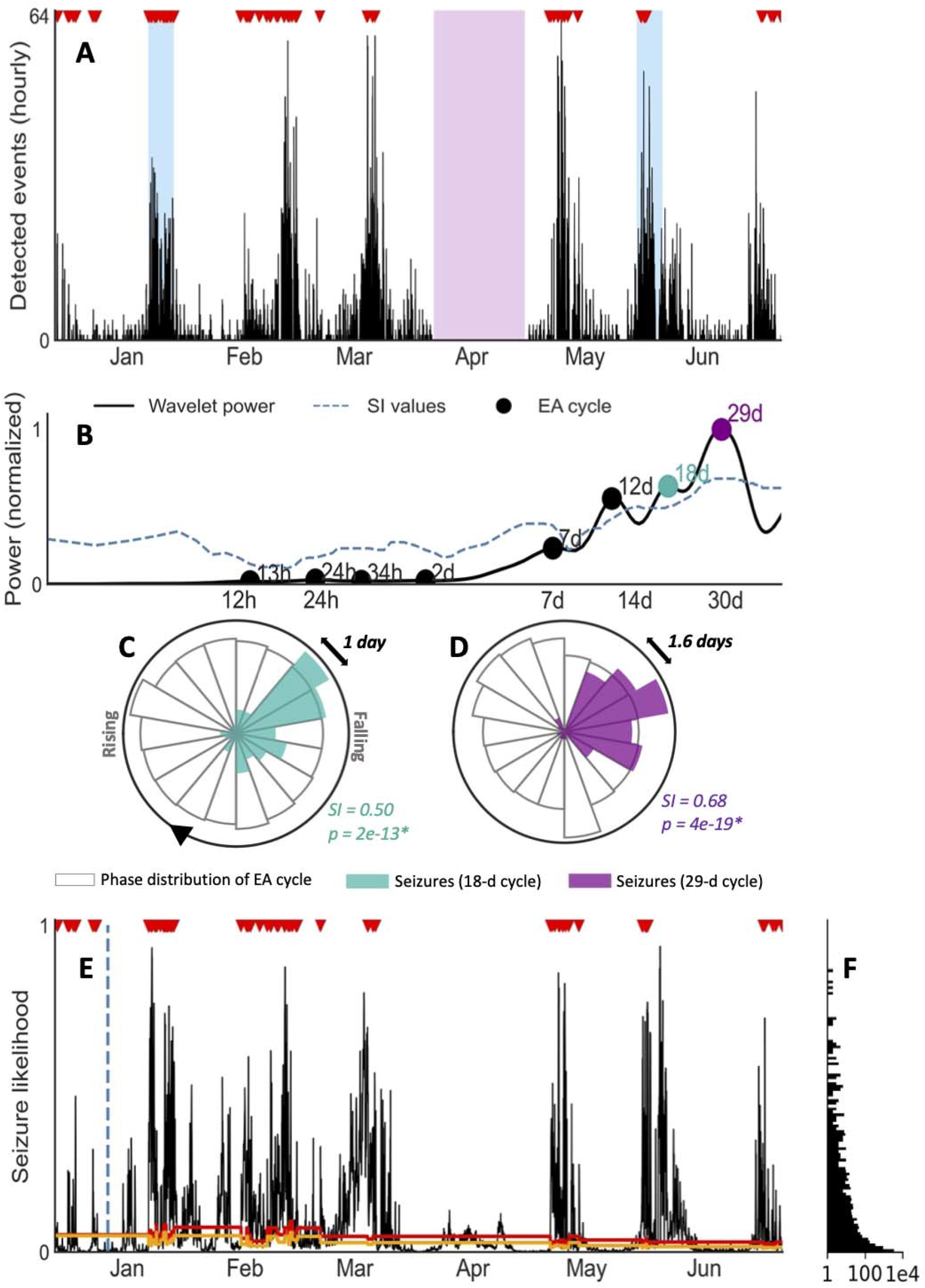
Seizure forecasting case study for patient 1. **(A)** Hourly rate of EA detected in the sub-scalp EEG device. Verified seizures (red markers) from review of sub-scalp EEG device are shown. The blue regions represent scalp video-EEG assessment periods, and the purple region represents a period where the device was turned off. **(B)** Cycle detection in EA using a Morlet wavelet approach. The wavelet spectrum is shown for a range of time periods (x-axis), with cycles reaching significance denoted by a black or coloured marker. The two coloured markers indicate the two cycles shown in the circular histograms (C,D). **(C**,**D)** Circular histograms showing the phase distribution of EA cycles (transparent bars) and seizure occurrence (coloured bars). Seizures were strongly locked onto 18 day (C) and monthly cycles (D) of EA. Seizures only occur in a narrow phase of the periodic activity suggesting a strong relationship between the cycle and seizures (p<0.05 with Omnibus test and high SI values). **(E)** Hourly likelihood of seizures. The likelihood of seizures occurring within the next hour is given by the black line and seizures are shown by the red markers. The training cut-off date (day 14) is indicated by the blue dotted line. The orange and red lines represent the medium and high risk thresholds, respectively. **(F)** Frequency (x-axis, log scale) of each seizure likelihood value (y-axis, shared with Figure 3E).

Figure 3A highlights the presence of multi day cycles (approximately monthly). The hourly event count was used to identify significant periodic cycles ranging from 12 hours to 98 days, as shown in the wavelet spectrum in Figure 3B. Seizures were only phased locked to some of these cycles (quantified by significant SI values). For participant 1, phase locking was found between seizures and cycles of 18 days and 29 days (Figures 3C,D).

A practical forecaster minimises the amount of time the forecaster displays a high risk warning while maximizing the number of seizures occurring during high risk. Alternatively, an opposite, suboptimal forecaster would always show high risk, achieving perfect predictive performance but of no utility to the end-user. Figure 3E demonstrates seizure likelihood over 6 months in participant 1, where risk levels have been optimised to be of highest utility to the user. The likelihood trace peaks in a cyclical manner, with seizures typically occurring close to the peaks. The participant had 134 seizures during this period, 15 of which occurred in the first 14 days (initial training phase) and 119 of which occurred during the testing period.

The time spent in low, medium, and high risk warnings and seizures that occurred during these periods are given in Table 3 for the testing phase. The forecast resulted in the participant spending 26% of time in a high risk state, 11% of time in a medium risk state and 63% of time in a low risk state. Of 119 testing seizures, 99 (83%) occurred during high risk, 8 (7%) occurred during medium risk and 12 (10%) occurred during low risk. The Area Under the Receiver Operating Characteristic Curve (AUC score), which demonstrates how good the model is at distinguishing non-seizure hours from seizure hours during testing, was 0.88. These results underscore the feasibility of seizure forecasting using data from the sub-scalp EEG device.

**Table 3.** Forecast results based on electrographic seizures. Number of electrographic seizures occurring in, and time spent in high, medium, and low risk states during testing. Training was performed on electrographic seizures. AUC = 0.88

|  | In high risk state | In medium risk state | In low risk state |
| --- | --- | --- | --- |
| **Seizures** | 99 (83%) | 8 (7%) | 12 (10%) |
| **Time** | 26% | 11% | 63% |

## Discussion

Here, we have successfully shown that a sub-scalp system can accurately record ultra-long term EEG (>12 months) and detect focal seizure activity (Figure 2). The device was well tolerated in all five participants, with no serious adverse events to date. This suggests that continuous monitoring of EA chronically is possible with a minimally invasive and discrete device. The benefits of this are ubiquitous, not only for seizure forecasting, but also for medication management, anti-seizure medication trials and surgical planning. It is highly likely that accurate and objective quantification of seizures and EA will become essential for future drug trials to provide more objective assessments of therapeutic benefit; sub-scalp EEG would be highly suitable for this purpose.

Our results demonstrate that sub-scalp devices record high quality neurological signals that are similar to scalp EEG. Sub-scalp recordings are also sensitive to other small neurological events such as sleep transients (Figure 2B). Lack of sleep and deviations from normal sleep patterns are known risk factors for seizures. Conversely, the treatment of seizures and seizures themselves can disrupt normal sleep patterns (40,41). Sub-scalp devices provide an opportunity to investigate the complex relationship between sleep and seizures and can aid in patient management and seizure forecasting (39). The sub-scalp EEG was less noisy compared to scalp EEG. Sub-scalp EEG contained much less interference from electrical line noise (50 Hz in Australia) and was not affected by movement artefacts typically observed in scalp EEG due to the movement of wires. Sub-scalp devices are, however, susceptible to other noise and artefacts, such as muscle activity recorded by electromyography (EMG). While jaw EMG artefacts may obscure the underlying EEG activity (Figure 2C), it can also identify jaw activity that is a feature of the seizures. In contrast, blinking artefacts could not be seen in the sub-scalp recordings, most probably because of the parietal positioning of electrodes (Figure 2D).

The sub-scalp device can be used to continuously monitor interictal and ictal events, which may provide better understanding of the burden of disease. This information is also of importance for clinical trials of novel therapies and for routine patient management. Currently, clinical trials of novel therapies rely on patient seizure diaries. The current study confirms the unreliability of patient reported events (Table 4) in line with previous findings (11). Our results highlight the large number of missed events (Table 2; participant 1 reported 39 seizures versus 134 actual seizures over 6 months), which impacts the estimate of disease burden and distorts the estimated benefit of new therapies. Our case study also highlights the long-term fluctuations that occur in the EA rates (Figure 3A), which have been implicated in cognition and memory performance (5,6). Understanding how EA changes is important for tailoring treatments that not only reduce seizures but ultimately improve quality of life.

The Minder™ sub-scalp system demonstrated utility in capturing cycles of EA. In the current work, there was clear rhythmicity in the EA (Figure 3A). This is concordant with previous work with invasive EEG showing the prevalence of circadian and multiday cycles in interictal EA (17,18) and seizures (20,29,31). Using a similar approach to previous work (20,40), cycles were detected at circadian and multiday periodicities for one individual (Figure 3B), with 18-day and 29-day cycles in EA showing the strongest relationship with seizure timing (Figure 3C,D). Interestingly, multiday cycles of EA in this subject were stronger than the circadian rhythm. Capturing multiday cycles requires long term monitoring and, in addition to demonstrated utility for forecasting, an understanding of seizure cycles may be critical for the development of new therapies.

We have also demonstrated the potential for seizure forecasting with sub-scalp systems. In this example, a forecaster achieved high accuracy (83%) with little time in a high risk state (26%), despite the high frequency of seizures in this participant. The AUC score (0.88) was comparable to recent, state-of-the-art forecasting using EA cycles derived from intracranial EEG (26,32). In contrast, a forecaster based on seizures reported by participant 1 was highly unreliable at predicting electrographic seizures (Table 4).

The case study demonstrates the high performance that can be achieved through an event-based seizure forecaster. This forecaster may be used to generate powerful prior probabilities for a more advanced seizure forecaster that combines other features, such as non-invasive information (e.g., medication adherence, heart rate etc.) and continuous features derived from the EEG (e.g., spectral power, autocorrelation etc.). Additionally, the forecast was able to continue making predictions despite missing EA during the period data was not collected. Whilst cycles in EA were attenuated during this period, seizure cycles were still utilised, as they rely on a fitted sinusoid of fixed period-length. The relative low likelihood of seizures during April compared to other months suggests that cycles in EA were stronger predictors of seizures than seizure cycles. This is in line with previous work, which suggests seizures are more robustly synchronised to cycles of a continuous biomarker than fitted sinusoids of fixed period-length (41).

Cyclic features in the EEG were stable and no adaptation period was required to start forecasting in this participant. The lack of implantation effect of sub-scalp systems (42) is in contrast to intracranial devices, which require a craniotomy and often significant period before the signal stabilises (21). This may require months of data to be discarded prior to training forecasting algorithms (28,32). In this case, forecaster training was undertaken immediately, and only 14 days of training data was required to generate forecasts (although this will depend on seizure frequency). Further work will investigate the utility of forecasting using sub-scalp recordings in a prospective study.

There are limitations with sub-scalp EEG systems. First, despite the limited invasiveness of subcutaneous electrodes, this surgical procedure may not be acceptable to all people with epilepsy (43). Hence, patient seizure diaries will remain a useful tool in clinical settings, and non-invasive forecasting systems based on mobile and wearable devices are desired by the epilepsy community (43,44). Wearable sensors and non-invasive features may be useful to forecast seizure likelihood (25,45), and self-reported events and biomarkers derived from wearables also demonstrate cycles that are co-modulated with seizure likelihood (30,38,40). However, the correlation between self-reported events and electrographic events is patient-specific. In cases where the accuracy is less than perfect, such as participant 1, it is unlikely that forecasts using self-reported events will perform as well as forecasts using chronic EEG. Despite advances in wearable technology for seizure detection, there remain significant false positives and many seizure types are missed (46). It is likely that chronic sub-scalp EEG recordings will prove to be a critical ‘ground-truth’ to develop wearable seizure detection and forecasting.

Second, validating electrographic seizures also remains a significant challenge, even with the aid of an algorithm detecting suspect events. A short 24-hour segment of continuous EEG alone can take hours for a trained neurophysiologist to review, which is not viable for large scale use of sub-scalp devices, and so optimising seizure detection algorithms will be critical.

This study has demonstrated the feasibility of using a continuous sub-scalp EEG device to record data of sufficient resolution to capture relevant events, detect the events algorithmically, and use the events in a seizure forecasting algorithm. This data is extremely valuable for the assessment of epilepsy and could be linked to systems to improve safety and independence, potentially changing fundamentally our approach to the management of the condition.

## Data Availability

Data was collected as part of an ongoing clinical trial. Following completion of the trial, data can be made available upon reasonable request to the corresponding author (MJC).

